# Intelligent Pneumonia Identification from Chest X-Rays: A Systematic Literature Review

**DOI:** 10.1101/2020.07.09.20150342

**Authors:** Wasif Khan, Nazar Zaki, Luqman Ali

## Abstract

Chest radiography is an important diagnostic tool for chest-related diseases. Medical imaging research is currently embracing the automatic detection techniques used in computer vision. Over the past decade, Deep Learning techniques have shown an enormous breakthrough in the field of medical diagnostics. Various automated systems have been proposed for the rapid detection of pneumonia on chest x-rays images Although such detection algorithms are many and varied, they have not been summarized into a review that would assist practitioners in selecting the best methods from a real-time perspective, perceiving the available datasets, and understanding the currently achieved results in this domain. This paper overviews the current literature on pneumonia identification from chest x-ray images. After summarizing the topic, the review analyzes the usability, goodness factors, and computational complexities of the algorithms that implement these techniques. It also discusses the quality, usability, and size of the available datasets, and ways of coping with unbalanced datasets.

## 1. INTRODUCTION

The chest X-ray (CXR) is an easy, economical, and commonly adopted tool for diagnosing lung diseases [1]. An experienced radiologist interprets an X-ray as either normal or presenting a disease such as lung cancer, tuberculosis, or pneumonia. One of the most common chest diseases is pneumonia, a lung infection caused by viruses, bacteria, or fungi [16]. Pneumonia is life threatening to infants, older adults, patients placed on a ventilator in hospital, and asthma patients. Moreover, pneumonia is a high-risk illness, especially in developing countries where millions of people are impoverished and lack access to medical facilities. The World Health Organization (WHO) [3] estimates that each year, over four million deaths are caused by pneumonia and other air pollution-associated diseases [4]. Over 150 million people, mainly children under five years old, are infected with pneumonia annually [5]. Viral pneumonia tends to be slight while bacterial pneumonia is more severe, especially in children [6]. The fungal type can occur in patients with weak immune systems. As CXRs are relatively low cost, they are more commonly requested than other medical modalities such as magnetic resonance imaging (MRI) and computed tomography (CT) [7]. The demand for CXRs translates to thousands of readings per radiologist per year; accordingly, there is a shortage of radiologists in both developing and developed countries [8].

Accurate and timely diagnosis is essential for reducing the mortality of lung diseases. In developing countries, where diagnoses and treatment are delayed by the shortage of knowledgeable radiologists, pneumonia in children is associated with alarming death rates. The massive gap between the number of doctors and the population of a specific area also hinders a timely diagnosis. Furthermore, CXRs have lower resolution than MRI and CT, and are not easily interpreted even by experienced radiologists. Decision making by medical staff can be supplemented by computer-aided diagnostic (CAD) tools, which combine aspects of computer vision and machine learning with radiological image analysis for recognizing and extracting patterns [9]. A typical CAD system sequentially processes the input data (CXRs), extracts the features, and classifies the features. The first step pre-processes the CXR data, the second step extracts the features from the input images by various techniques such as Gaussian filters [10], edge detection [11], and morphological operation [12], and the third stage distinguishes the extracted features by a suitable classifier such as a Support Vector Machine (SVM) [13], Random Forest (RF) [14], or a Neural Network algorithm [15].

Publicly available CXR datasets for pneumonia are highly class imbalanced, meaning that more images are available in one class than in other. Class imbalance seriously degrades the efficiency of a classification system. For instance, the dataset compiled by Kermany et al. [16], which is used as an example in the present work, includes 5856, 4300 images in the disease (pneumonia) class and 1583 images in the normal class. Class imbalance problems can be tackled by several pre-processing techniques. Traditionally, the minority class is over-sampled by duplicating randomly selected samples while the majority class is under-sampled. However, random over-sampling introduces the overfitting problem, and under-sampling loses valuable information. Also, the sampling technique itself has limited generalization and variance [17]. The training dataset is often expanded by data augmentation, which creates false representations of the original images to avoid overfitting [18]. More recently, artificial data have been generated by generative adversarial networks (GANs) [19, 54], which consists of two neural networks: a generator and a discriminator. The generator synthesizes artificial samples with the required variations from the input data distribution, and the discriminator differentiates between the samples generated by the generator and the samples in the input data. Thoroughly engineered architectures such as deep convolutional GAN (DCGAN) [2], styleGAN [21], and Cycle GAN [22] have been introduced for more robust synthesized data generation.

Traditional CAD-based systems [23] have successfully classified lung diseases in CXRs, but require extensive handcrafted techniques for feature-extraction from images, followed by machine learning classifiers [23], [52], [53]. Such techniques are affected by noise and may need to be adapted to different problems. To overcome these problems, researchers have developed many artificial intelligence (AI) based solutions. Data-driven deep learning (DL) methods achieve automatic end-to-end feature extraction and classification. The convolutional neural network (CNN), a type of deep neural network, has achieved ground-breaking results in different tasks related to pattern recognition over the last decade. Inspired by the visual cortex of humans [27], CNNs differentiate a large number of classes in image recognition problems [18] [28]. However, they require a large volume of training data to learn the better feature patterns [29], and cannot easily obtain a large labeled and balanced dataset for medical applications. Medical image classification by a CNN is usually performed either by training the CNN from scratch, using an existing pre-trained network without retraining, or fine-tuning a pre-trained network on a target dataset [30].

Recently, DL-based algorithms have become the default choice for medical image applications. Examples are SegNet [31], DenseU-Net [32], Chexnet [33], and CardiacNet [34]. DL-based techniques are similarly applied to pneumonia detection in CXRs [35]. DL can also be implemented by techniques other than CNN, such as transfer learning. This paper presents the current literature on pneumonia identification in CXR images. Specifically, it analyzes the usability, goodness factors, and computational complexity of the algorithms that perform these techniques. It also discusses the quality, usability, size, and balance extent of the available datasets.

The remainder of the paper is organized as follows. Section II elaborates on the datasets and their respective details. Section III focuses on the data pre-processing and augmentation techniques that solve the unbalanced class problem, and Section IV reviews the techniques applied to lung disease detection. Section V describes the evaluation metrics and provides a brief comparative analysis. Section VI finalizes the discussion and draws the conclusions.

## 2. RESEARCH METHOD

A systematic literature review (SLR) is performed with a research method that must be unbiased and ensure completeness to evaluate all available research related to the respective field.

### 2.1. Data Sources

We used four electronic databases as primary data sources to search for the relevant studies. The electronic databases used in the search process are listed in Table 1.

**Table 1:** Data Sources

| Identifiers | Databases | URL | Results |
| --- | --- | --- | --- |
| ED1 | IEEE Xplore | https://ieeexplore.ieee.org/ | 74 |
| ED2 | Science Direct | https://www.sciencedirect.com/ | 80 |
| ED3 | Springer Link | https://link.springer.com/ | 74 |
| ED4 | ACM | https://dl.acm.org/ | 79 |

### 2.2. Search Terms

These are the search terms adopted from the research question and literature

- pneumonia detection OR secure OR Health Safety infectious dieses of chest OR x-ray dataset OR deep learning for pneumonia
- Chest radiography OR Chest x-rays OR Chest Disease detection
- data balancing AND chest X-rays OR pneumonia, Generative adversarial networks (GANs) AND chest X-rays
- Search string for the automated search is given below:

(Chest OR Pneumonia OR Diseases OR x-ray OR Xray OR CXR OR lung diseases)

### 2.3. Study selection procedure

#### 2.3.1. Inclusion and exclusion criteria

Inclusion and exclusion criteria are employed to extract relevant studies from different data sources in order to answer the research questions.

##### 1) Inclusion

- Studies related to Pneumonia Identification using chest x-ray.
- Studies that are not focused on pneumonia detection but add value to solve the problem.
- Studies that were published in a peer-reviewed conference or journal.
- Studies that were published in or after 2010 to early 2020.
- Studies in the English language.
- Studies that consist of peer-reviewed publications.

##### 2) Exclusion

- Studies other than Pneumonia Identification by using chest x-ray.
- Studies other than English.
- Studies with no validation of the proposed technique.
- Editorials, short papers, posters, technical reports, patents, reviews, blocks information, Wikipedia, survey, and extended papers.

## 3. DATASETS

### 3.1. THE CHEST X-RAYS 14 DATASET

Wang et al. [1] presented a chest X-ray database named Chest X-ray14 [1] [36], which contains 108,948 images of eight (now 14) diseases collected from 32,717 patients. This dataset was acquired from Picture Archiving and Communication Systems (PACS). The eight common thoracic diseases—Atelectasis, Effusion, Infiltration, Cardiomegaly, Mass, Nodule, Pneumonia, and Pneumothorax—were shortlisted as keywords in a search of the PACS system, and the matching reports were extracted along with their associated images. Examples of each lung disorder are shown in Fig. 1. The typical size of the x-ray images (3000 × 2000) pixels was resized to 1024 × 1024 pixels without losing any significant details or content. Among the 108,948 images, 24,636 images contained one or more pathologies, while the remaining 84,312 images were normal. Fewer than 1,500 pneumonia images were found in this dataset, which is publicly available.

**FIGURE 1.**
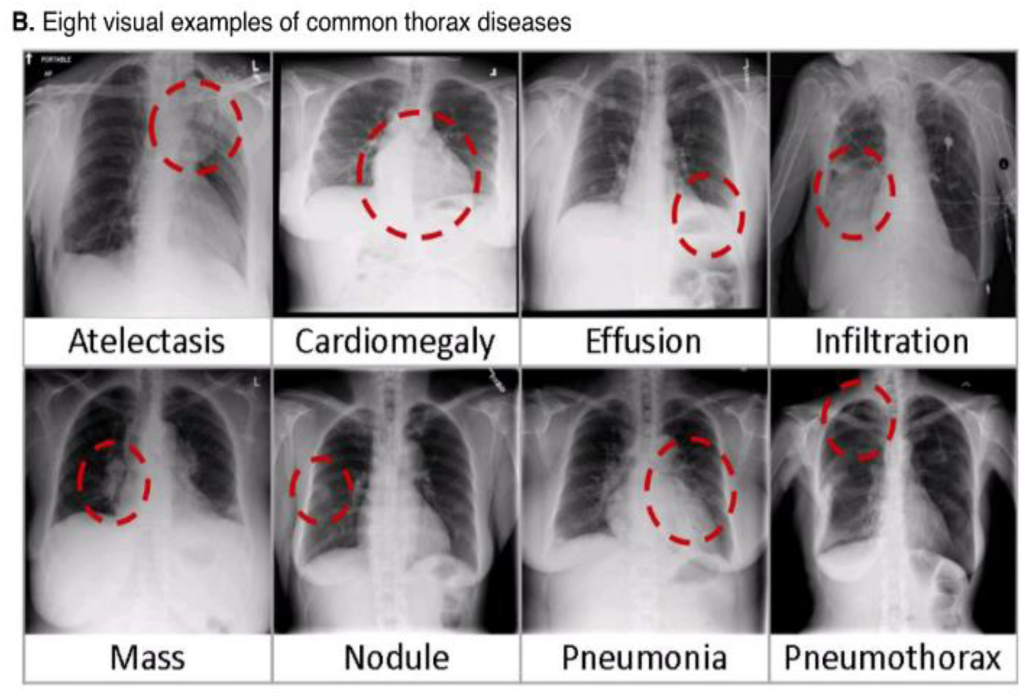
Examples of lung-disease images extracted from the Chest X-rays14 dataset.

### 3.2. PEDIATRIC CXRS FOR PNEUMONIA DETECTION

Kermany et al. [16] collected and labeled pediatric CXRs from the Guangzhou Women and Children’s Medical Centre (Guangzhou, China). All CXR imaging was performed as part of the patients’ routine clinical care. The CXR images for the present study were selected from pediatric patients aged 1 − 5 years, thus obtaining 5856 images including 3,883 pneumonia images (2538 bacterial, 1,345 viral) and 1,349 typical images. The x-ray images are available in various dimensions: 1040 × 664, 1224 × 1000, and 1848 × 1632. Figure 2 is an example taken from Kermany et al.’s dataset.

**FIGURE 2.**
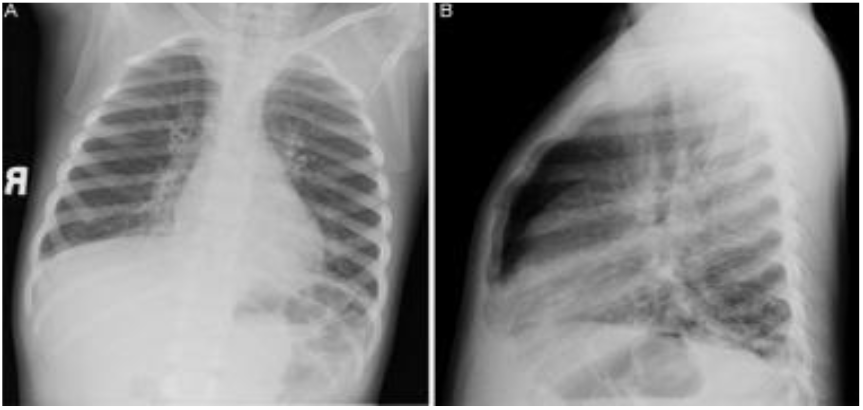
Sample taken from the Pediatric CXR dataset.

### 3.3. MIMIC CXR

MIMIC–CXR–JPG [37] is currently the largest publicly available CXR dataset worldwide, containing over 377110 CXRs associated with 227827 imaging studies. It contains the data of 14 chest diseases sourced from the Beth Israel Deaconess Medical Centre (Massachusetts, USA) [38] from 2011 to 2016. Examples of the images from this dataset are presented in Fig. 3.

**FIGURE 3.**
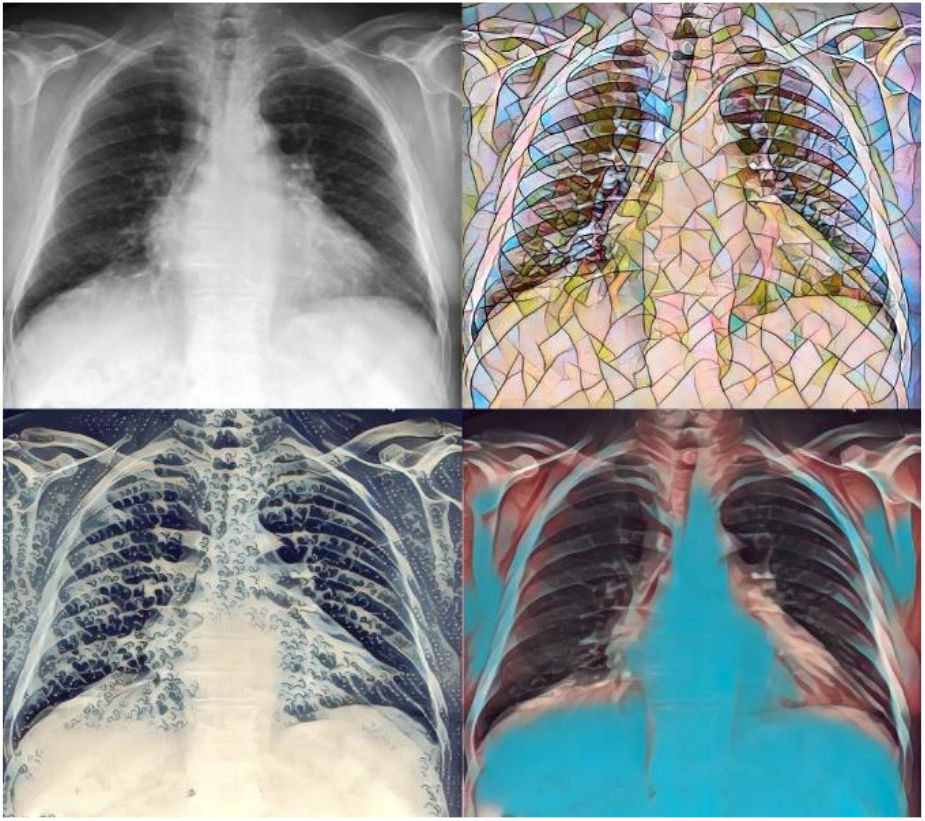
Sample images taken from the MIMIC dataset.

### 3.4. INDIANA DATASET

Demner-Fushman et al. [39] collected the Indiana (USA) dataset from multiple hospitals associated with the Indiana University School of Medicine. This dataset is composed of 7470 chest radiographs. The images are annotated with the disease view (frontal or lateral), and include pulmonary edema, cardiac hypertrophy, pleural effusion, and opacity. This dataset is publicly available for users but contains no pneumonia samples so cannot be used as a pneumonia detection dataset. Samples from this dataset are presented in Fig. 4.

**FIGURE 4.**
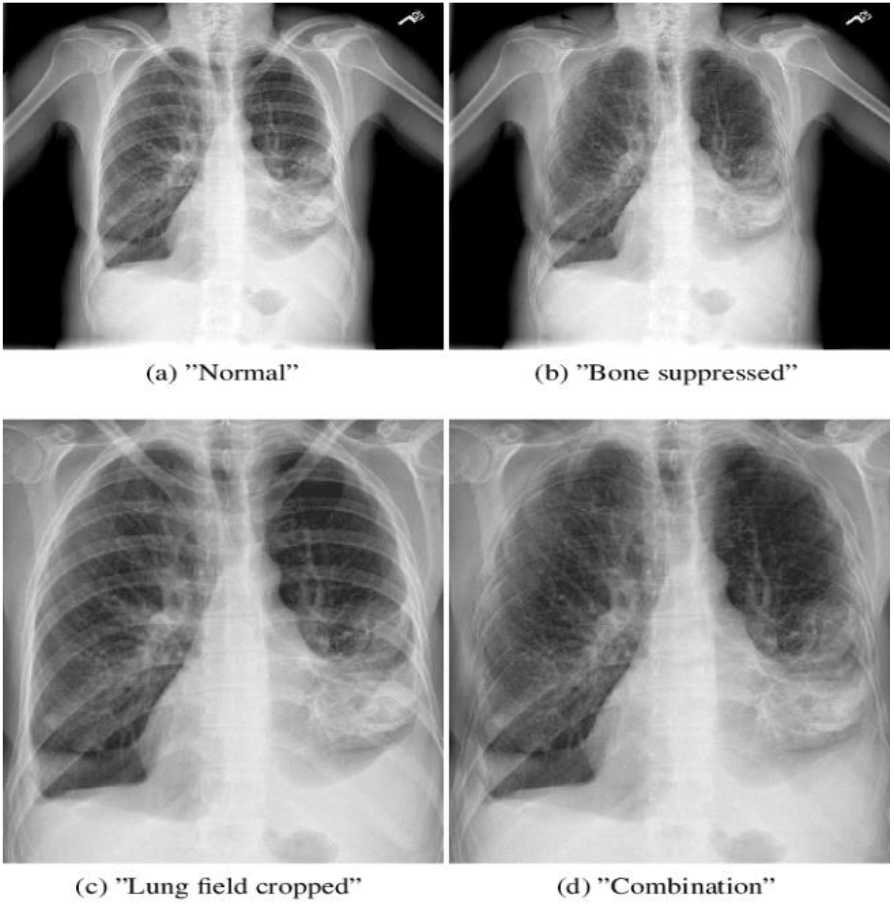
Sample images taken from the Indiana dataset.

### 3.5. MC DATASET

Jaeger et al. [40] gathered 138 chest images with the collaboration of Montgomery County’s screening program for tuberculosis (USA). Among these images, 80 were obtained from healthy subjects, and 58 were obtained from tuberculosis subjects. The resolution of the images varies from 4020 × 4892 to 4892 × 4020 pixels. The dataset is publicly available for users, but as it contains no pneumonia samples, it cannot be used as a pneumonia detection dataset. Typical images in the MC dataset are presented in Fig. 5.

**FIGURE 5.**
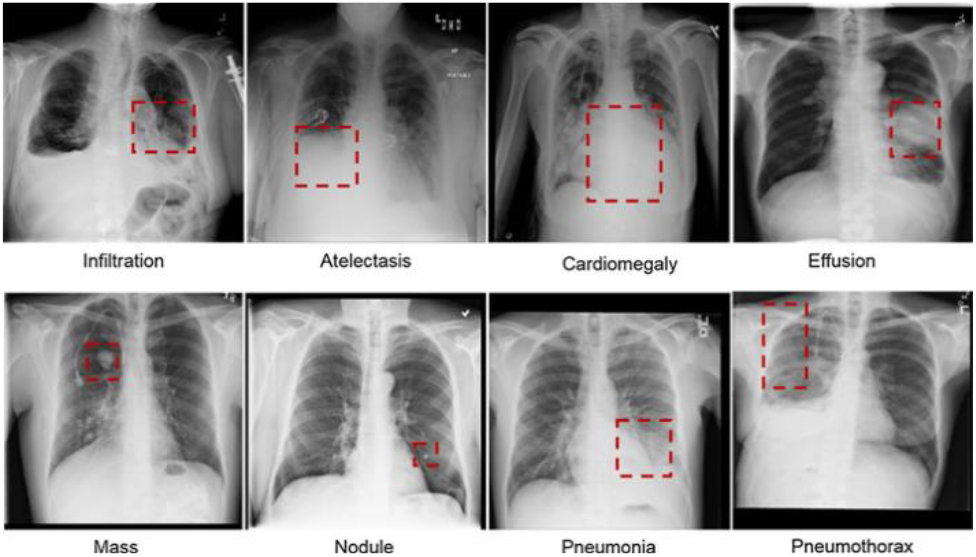
Sample images taken from the MC dataset.

### 3.6. SHENZHEN DATASET

In addition to the MC database, Jaeger et al. [40] collected the Shenzhen dataset from Shenzhen’s hospital (Guangdong Province, China), named the Guangdong Medical College. This dataset contains 662 chest radiographs: 326 healthy and 336 tuberculosis cases. Some examples are displayed in Fig. 6.

**FIGURE 6.**
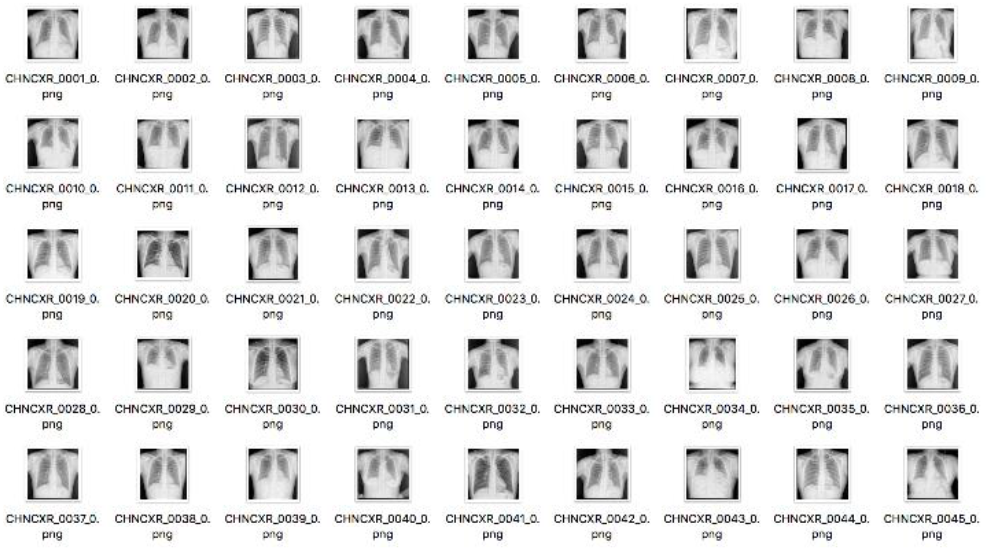
Sample images taken from the Shenzhen dataset.

### 3.7. KIT DATASET

Ryoo and Kim [41] collected 10848 images from the Korea Institute of Tuberculosis. This dataset includes 7020 images from healthy subjects and 3828 images from tuberculosis cases.

### 3.8. JSRT DATASET

The Japanese Society of Radiological Technology [42] collected 247 chest images, among which 154 presented with pulmonary nodules, and 93 were nodule-free. All X-ray images are sized 2048 × 2048 pixels, and the f depth of the grayscale is 12 bits. Examples are displayed in Fig. 7.

**FIGURE 7.**
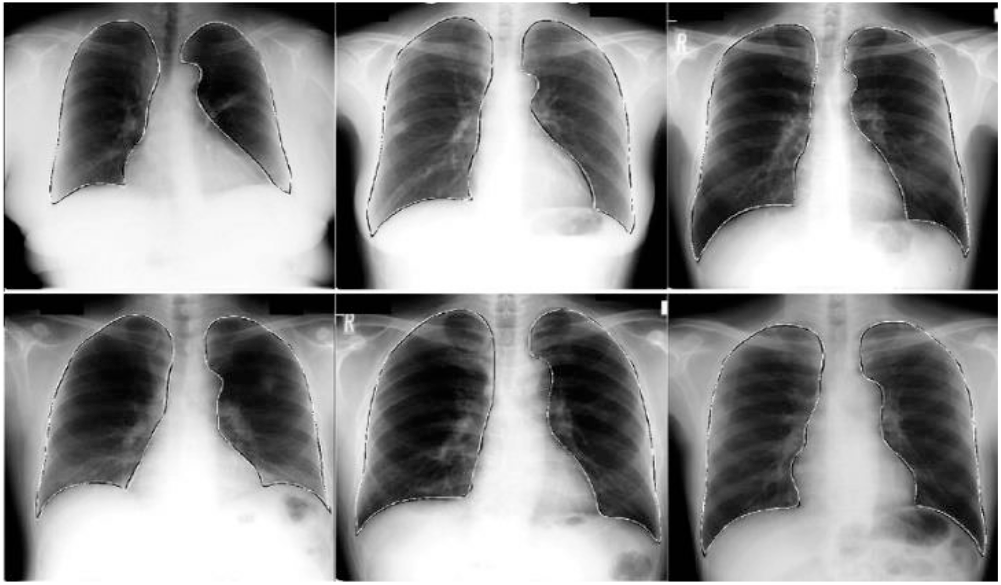
Sample images taken from the JSRT dataset.

## 4. DATA PRE-PROCESSING

Most CXRs are obtained in Digital Imaging and Communications in Medicine (DICOM) format with a large amount of metadata, but the DICOM format is difficult to understand by experts outside the radiology domain [37]. In other domains, the images are usually stored in formats such as PNG and JPEG, which are processed by compression algorithms to conserve the image information without losing any desired information. First, the patient’s information is de-identified to satisfy the mandatory privacy standards. This step requires the removal of the patient’s details, identifiers, and dates by a customized algorithm combining image processing and optical character recognition, which detects and removes text in the X-ray image. After de-identification, the DICOM images are converted into JPEG or bitmap format by the approaches described in [1][37].

Moreover, the dimensions of normal X-ray images (3000 × 2000 pixels) are difficult to handle by both deep CNNs and the computing hardware. Therefore, X-ray images must be reduced to an optimal size while preserving their vital information. Wang et al. [1] reduced the size of X-ray images to 1024 × 1024 pixels, open to 512 × 512 pixels. The sizes of the MIMIC [37] and paediatric CXR datasets are 2048 × 2048 and 1024 × 1024 pixels, respectively.

### 4.1. DATA BALANCING, AUGMENTATION, AND ENCANCEMENT BY TRADITIONAL TECHNIQUES

Most of the datasets containing pneumonia images are class-imbalanced, which biases the classification toward the majority class. The imbalance problem has traditionally been resolved by the following techniques.

The mean squared error (MSE) sums the errors in the whole dataset and calculates their average. The MSE-based loss function fails when the data are imbalanced, as it favors the majority. Wang et al. [43] proposed the mean false error (MFE) and its improved version, the mean squared false error (MSFE), for training neural networks on imbalanced data. This loss function computes the average errors on a class-by-class basis and adds them, ensuring that each class has the same loss value. Data sampling can also help in data balancing problems [35]. Samples can be randomly duplicated from the minority class and randomly removed from the majority class. However, as mentioned above, random duplication and under-sampling result in overfitting (owing to duplicate samples), and loss of important information, respectively. Under-sampling and over-sampling methodologies for solving data-unbalanced problems have been substantially developed since the late 1990s [45-53]. Numerous researchers have suggested and employed different sampling techniques.

Several reviews have focused on the effectiveness and usability of these methodologies [45], [[46]. Random sampling, as proposed by some researchers, has several shortcomings. First, it can eradicate important relevant samples from the data, thereby risking the overfitting problem. Kubat [47] selected samples from the original population by one-sided selection, forming an under-sampled dataset. To identify the bad samples in the data, the authors employed the Tomek links algorithm [48] and the condensed nearest neighbor method [49]. The latter algorithm filters out the noisy and bad examples from the majority class that needs to be under-sampled.

Laurikkala [50] suggested the Neighborhood Cleaning Rule for filtering excess examples from the majority class data. They calculated three nearest neighbors for every example (*D*_*i*_) from the training set such that if *D*_*i*_ fits within the common class but has been misclassified by the chosen nearest neighbors, it (sample) is detached from the dataset. Similarly, if *D*_*i*_ locates within the less common class and has been misclassified by the chosen nearest neighbors, it should be detached from dataset. This approach can hit a computational bottleneck when processing large, highly imbalanced datasets. The synthetic minority oversampling technique (SMOTE) [51] creates simulated data based on the similarities between pairs of existing minority samples. However, the SMOTE technique is limited in generalization and variance.

Wang [52] incorporated SMOTE into a locally linear embedding algorithm (LLE) that maps high-dimensional data into a low dimension. They collected three datasets of CXR images and checked the pulmonary detection performances of their method using multiple classifiers (k-neural network, SVM, and naïve Bayes). The minority-to-majority class ratio was almost 1:25. In terms of classification accuracy, the LLE–SMOTE algorithm outperformed conventional SMOTE by 2–4%. Krawczyk et al. [53] presented several data sampling techniques for class-imbalanced problems (SMOTEBoost, OverBagging, RAMO, ADASYN, and conventional SMOTE) and tested them on a dataset of 340 images for breast cancer detection. The majority class contained 144 images of intermediate malignancy, while the minority class contained 26 images of high malignancy (giving an approximate imbalance ratio of 6:1). As the base classifier, they employed an SVM with a Gaussian kernel and a minimal optimization training procedure. SMOTEBoost achieved the highest sensitivity (88.46%) and specificity (exact negative rate = 88.8%) among the tested oversampling methods.

### 4.2. DATA BALANCING USING GENERATIVE ADVERSARIAL NETWORKS (GANS)

First introduced by Goodfellow et al. [54], a GAN generates new samples based on the learned input (training) data distribution *p*. GANs consist of two multi-layer perceptrons: a discriminator (*D*) and a generator (*G*). The generator creates samples from a simple distribution *p*(*g*) such that *p*(*data*) *= p*(*g*). During the training process, *G* maximizes the probability of *D* making an error, and creates fake samples that follow the input data distribution [5]. After several steps of training, the goal *p*(*data*) *= p*(*g*) is achieved and *D* cannot discriminate between real samples and samples from *G*. In this way, the GAN generates synthetic samples. GANs have attracted attention for their high performance but are unstable during training, often producing nonsensical outputs from *G* [54]. To overcome these problems, researchers have recently developed modified GANs such as DCGAN [2], styleGAN [21], and Cycle GAN [22].

Wei et al. [55] proposed the structure-correcting adversarial network (SCAN) for organ segmentation (left lung, right lung, heart) in CXRs. SCAN is suitable for small training datasets. The algorithm was evaluated on the publicly available JSRT [42] [56] and Montgomery [40] [57] datasets. They trained a fully convolutional network (FCN) on a small dataset of grayscale CXR images. The FCN optimized the segmentation network, whereas the SCAN differentiated the segmentation network predictions from the ground truth annotations. Data augmentation was not applied, as it did not improve the results. The algorithm was tested by the intersection-over-union (IoU) and Dice coefficient metrics. In the JSRT evaluation, the adversarial training improved the performance from 92.9% in FCN to 94.7% in the proposed algorithm.

Adar et al. [58] isolated the region of interest (ROI) in a synthetic liver lesion using a GAN architecture, and classified its features using a CNN. They used a limited dataset of 182 CT images of liver lesions (53 cysts, 64 metastases, and 65 hemangiomas). The sensitivity and specificity were improved from 78.6% and 88.4% respectively in classic data augmentation to 85.7% and 92.4% respectively in the GAN-generated synthetic data augmentation. However, the ROI segmented in their paper was excised only from a 2D CT image. Gupta et al. [25] proposed a GAN for bone-lesion detection. To reduce the computational resources, they extracted a lesion patch from the X-ray rather than from the full image, and thereby obtained many patches. For training, the lesions were annotated manually under expert advice. A non-lesion patch was translated into a lesion patch.

Salehinejad et al. [20] proposed DCGAN for generating artificial CXR images from real x ray images. The dataset was obtained from the Radiology Information System, which stores the data of *four* diseases, and the classification was performed by AlexNet [18]. The accuracy improved from 88.4% in [58] to 92.1% in DCGAN, but the generated images had relatively low resolution. Chuquicusma et al. [59] artificially generated CT images of lung nodules by DCGAN, and checked their quality in a visual Turing test by two radiologists. If the radiologists confused the original images with the DCGAN-generated samples, then the DCGAN was judged to produce highly realistic samples.

Chuquicusma et al. [59] extracted 145 lung nodules (635 benign, and 510 malignant) from the Lung Image Database Consortium image collection, and supervised a visual Turing test by two radiologists, one with 13 years of experience (Radiologist 1), the other with four years of experience (Radiologist 2). Radiologist 1 correctly assessed 67% and 58% of the fake and real generated nodules, respectively. Meanwhile, Radiologist 2 correctly evaluated 100% and 92% of the fake and real nodules, respectively. The inter-observer agreements of the benign and malignant real cases were 44.91% and 58.56%, respectively. This experiment confirmed that DCGAN generates realistic samples. However, some samples contained the signs of both benign and malignant nodules, which were confusing and inferior in quality.

Baur et al. [60] proposed the deeply discriminated GAN (DDGAN) algorithm, a modified Laplacian pyramid GAN (LAPGAN) that generates realistic high-resolution samples of skin lesions from a set of 2000 training samples. The algorithm was evaluated on the 2017 International Skin Imaging Collaboration (ISIC) [61] dataset, which contains 2000 samples (374 melanomas, 1372 benign lesions, and 254 seborrheic keratosis samples). The performance of LAPGAN was compared with those of DCGAN and DDGAN. Although LAPGAN slightly outperformed DDGAN, it proved challenging to train, and its hyperparameters were difficult to adjust. Meanwhile, when trained on 374 images, DCGAN failed but DDGAN trained easily with fast convergence and no degradation from severe high-frequency artifacts; accordingly, it realistically generated high-quality images of 256 × 256 pixels.

Malygina et al. [62] attempted to solve the unbalanced data problem using CycleGAN for sample creation and DenseNet for classification. Pollastri et al. [63] modified the LAPGAN and DCGAN algorithms to generate skin lesion samples and their segmentation masks, which ease the task of adding data to a training set. After training, LAPGAN and DCGAN generated 192 × 256 pixel-sized images from the ISIC training dataset, and their associated binary segmentation masks. Data augmentation was performed on both real and generated images. In various experiments, LAPGAN outperformed DCGAN when trained on only 500 annotated images, but DCGAN’s performance rose when the number of annotated samples increased.

Using a conditional GAN and an active learning technique, Mahapatra et al. [64] generated CXR images from the Segmentation in Chest Radiographs (SCR) database, which contains 247 lung images (153 nodules and 93 healthy). They varied the number of samples in the initial training set, and showed that even when trained and tested on 35% of the full dataset, the method achieved almost the same results as fully supervised learning (training and testing on the whole dataset). An image and its manually segmented mask were input to the sample generator to generate realistic-looking images. After their informativeness was checked by a Bayesian Neural Network (BNN), the generated images were added to the training data to fine-tune the classifier. The test set included 400 images (200 normal, 200 nodular). When trained on 35% of the labeled data, the accuracy was 91.9%, versus 92.4% in fully supervised learning.

## 5. DATA MINING TECHNIQUES

### 5.1. TRADITIONAL MACHINE LEARNING

Oliveria et al. [65] proposed a machine learning-based network that classifies pediatric CXR images as pneumonia or normal images. They constructed an up-to-date database of 40 images (20 with pneumonia, 20 without pneumonia), and evaluated the system on 20 random test images from the database. Among various wavelet coefficients, the Haar wavelet transforms provided the highest accuracy of feature extraction from x-rays (97%), but a mediocre specificity (80%). Macedo et al. [66] proposed a system that detects pneumonia from chest radiographs by machine learning algorithms with the Haar wavelet transform as the feature extractor and a *k*-nearest neighbor (KNN) classifier. The model was trained and evaluated on a dataset of 166 child chest radiographs verified by a radiologist.

Sousa et al. [67] compared the performances of different machine learning algorithms in childhood pneumonia detection from CXRs. They utilized their self-generated pneumonia-detection dataset called PneumoCAD-dataset, which contains 156 grayscale chest radiographs. These images were first annotated by expert radiologists following the WHO guidelines. The authors extracted several texture-based features for the classification task: the coefficient of variation, energy, contrast, average energy, correlation, entropy, difference variance, average deviation, difference entropy, inverse, difference moment, sum average, residual mean, sum entropy, variance, sum variance, standard deviation, and suavity [75-78]. The SVM, KNN, and naïve Bayes classifiers on these extracted features achieved accuracies of 77%, 70%, and 68%, respectively.

Sousa et al. [98] proposed a pneumonia detection algorithm and tried five machine learning classifiers (KNN, naive Bayes, multi-layer perceptron, decision tree and SVM) combined with different dimensionality-reduction techniques (principle component analysis (PCA), sequential forward elimination, and kernel-PCA). For each combination, they tested the pneumonia-detection performance of their algorithm on CXRs. The dataset and features were those used in [23]. When 13-dimensional features produced by the kernel PCA were classified by naïve Bayes, their algorithm achieved an accuracy of 96%.

Depeursinge et al. [68] similarly compared the performances of five machine learning classifiers (naïve Bayes, KNN, J48 decision trees, multi-layer perceptrons, and SVM) in pneumonia detection. Their self-collected dataset contained 843 radiographs with their annotated ROIs. They extracted a total of 39 texture-based attributes and optimized the parameters of each classifier by grid-searching. The performance was evaluated by McNemar’s statistical tests and the accuracy measure. They concluded that SVM achieved the best values of each metric, with a correct prediction rate of 88.3%.

Yao et al. [69] proposed a machine learning-based automated system that identifies five diseases, including pneumonia. They collocated 40 CXRs and employed SVM classifiers with texture-analysis ability. Their extracted feature vector contained 25 texture features, such as the mean, variance, energy, and correlation. The co-occurrence matrix was also calculated. They achieved an accuracy of 80% in a pneumonia detection task.

Naydenova et al. [70] employed machine learning methods in a novel diagnostic process. Their methods input numerous clinical measurements that can be calculated by cheap and easy utensils. To evaluate their findings, they collected a dataset of 1093 patients, including 777 pneumonia samples and 316 healthy samples. They also extracted 47 clinical characteristics tools and employed nine feature-selection techniques (correlation coefficients, minimum redundancy, Gram– Schmidt, Relief, orthogonalization, selection operator, least angle shrinkage, elastic net, and sparse linear discriminant analysis). The extracted features were learned by the SVM and RF machine learning procedures. The authors also employed a predictive algorithm with four predictive attributes (temperature, heart rate, oxygen saturation, and respiratory rate). They reported a sensitivity of 96.6%, a specificity of 96.4%, and an area under curve (AUC) of 97.8%.

Antin et al. [71] proposed a supervised learning technique for pneumonia detection in CXR images. They accessed the NIH database of 112120 radiographs collected from 30805 patients. These images are preannotated with “one”, “more than one” or “no” diseases by expert radiologists. The authors selected 500 random samples and colored them by *k*-means clustering. The colored samples were preserved as the ground truth set. The classification results of a supervised learning methodology were verified by logistic regression. The authors claimed comparable performances in their experiments and discussed how deep learning could improve the outcomes of this task.

### 5.2. DEEP LEARNING

A CNN consists of three main parts: the input images, an in-depth feature extractor, and a classifier. Through its multiple layers, the feature extractor automatically learns the essential features from the raw input (or pre-processed) images. The learned features are passed to a classifier such as SoftMax, where they are classified based on the learned features. A CNN contains several layers, namely, a convolutional layer, a pooling layer, an activation layer, a dropout layer, and the classifier (e.g., SoftMax) [72]. A CNN can be built from scratch [6], by employing an existing pre-trained network without retraining, or by fine-tuning a pre-trained network on a target dataset.

### 5.3. TRANSFER LEARNING (OF PRE-TRAINED NETWORKS)

Wang et al. [1] classified pathologies from CXRs using various pre-trained models (AlexNet [18], GoogleNet [74], VGG16, and ResNet). The ResNet model (AUC = 0.63) proved more accurate than the other pre-trained networks. Yao et al. [77] proposed a two-stage end-to-end model with a DCNN encoder and an RNN (long short-term memory) decoder that predicts the labels of the pathologies in Chest X-ray14 data. The DCNN was a modified DenseNet trained from scratch on the Chest X-ray14 dataset. The classification accuracy was 71.0%. To improve this result, Rajpurkar et al. [33] proposed Chexnet, a pre-trained 121-layer DenseNet trained on the Chest X-ray14 dataset. The Chexnet algorithm achieved a higher performance of the F1 metric than experienced radiologists. For pneumonia detection, the dataset was divided into training (98637 images), validation (6351 images), and testing (420 images) sets. To apply the algorithm to all 14 diseases in Chest X-ray14, the dataset was also split into a 70%: 10%: 20% ratio of training, validation, and test sets. The accuracy of pneumonia detection was 76%.

Kermany et al. [16] proposed a transfer learning algorithm for retinal OCT diagnosis based on transfer learning of retinal OCT images, and reported state-of-the-art performance. The same transfer learning system was applied to pediatric chest x rays [16]. In normal versus pneumonia images, the accuracy, specificity, and sensitivity of this system were 92.8%, 0.90 and 0.93, respectively. Meanwhile, the accuracy, specificity, and sensitivity of detecting bacterial versus viral pneumonias were 90.7%, 0.88 and 0.90, respectively. Ayan et al. [73] compared the performances of two pre-trained networks (VGG16 and Xception [78]) on the same dataset. The sensitivity and specificity of distinguishing normal from pneumonia images were 0.82 and 0.91 respectively for VGG16, and 0.85 and 0.76 respectively for Xception.

Togacar et al. [79] applied three deep learning methods (Alexnet, VGG16, and VGG19) in a deep feature learning model that extracts CXR images at the eighth fully connected layer. The 1000 features obtained from each deep learning model were reduced to 100 by minimum redundancy maximum relevance. Finally, the selected features were combined into a latent Dirichlet allocation model, which achieved a 99% classification accuracy on a CXR dataset [16]. However, the authors did not apply the standard data division given in [16]; instead, they balanced only the normal images by data augmentation. Similrly, Liang et. al. [102] proposed 49 layers convolutional layers residual structure that overcome overfitting for pneumonia detection on pediatric chest X-rays dataset [16].

Putha et al. [80] trained a deep learning system on the largest available dataset, which contains 2.3 million CXR images with different pathologies. The deep learning algorithm (ResNet) was validated on 100000 of these images (CQ100k) and was further evaluated on 2000 images (CQ2000) collected from three hospitals in India. The pathologies were obtained from the images and their associated reports by a natural language programming (NLP) algorithm. To evaluate the correctness of the NLP algorithm, the 1930 x-ray images were independently verified by experienced radiologists. The accuracies of pathology classification on the CQ2000 dataset ranged from 89.0 to 99.0%. Similar results were obtained on the CQ100k dataset, but the accuracy of detecting normal x-ray images in this dataset reduced to 86.0%.

### 5.4. DL and non-image features

Baltruschat et al. [35] supplemented the CXR dataset with the non-image features (patient age, gender, and acquisition type). The authors applied the transferred learning technique with and without fine-tuning, as well as a CNN trained from scratch. The data were split in two ways. In the first data splitting, patients with multiple follow-up records were assigned to a single subset. This splitting enables a vast range of patient numbers; for example, 22420 images can be split into 5817 patients (split 2) or 6245 patients (split 5). The second dataset split is described in Wang et al. [1]. The former split improved the accuracy of classifying pneumonia by transferred learning from 75.3% to 76.7 % (for image dimensions of 448 × 448 pixels. The latter split reduced the AUC result of detecting pneumonia with ResNet-38 to 0.71, but achieved superior results on the five other diseases in the CXR dataset.

Bar et al. [82] concatenated the Decaf [26] features with the low-level features (the GIST [83] and bag-of-words features), and classified lung pathologies by a feature selection method. For feature selection, they selected the 5000 most significant features among the 18920 features obtained by concatenating the Decaf and low-level features.

Er et al. [44] performed a comparative study of different neural networks in chest disease diagnosis. Their dataset was obtained from Diyarbakir Chest Diseases Hospital from southeast of Turkey hospital in Turkey, which contains the epicrisis reports of patients with tuberculosis (50 samples), COPD (71), pneumonia (60), asthma (44), or lung cancer (32), along with healthy samples (100). The authors collected 357 samples and analyzed 38 non-image features, including complaints of coughing, weakness, chest aches, and high body temperature. In the pneumonia analysis (60 samples), the multilayer neural network with the Levenberg–Marquart architecture (one hidden layer) achieved the highest accuracy for pneumonia detection (91.67%). No other metrics were presented in their paper.

### 5.5. Customized CNN

Stephan et al. [5] proposed a CNN with four convolution layers, which they trained from scratch on the pediatric CXR dataset proposed by Kermany [16]. They split the data differently from Kermany, assigning 2134 images to the validation set, and increasing the training dataset by augmentation techniques. They reported an accuracy of 93.7% on the validation set, but did not consolidate this performance by other metrics. This lack of further verification is a significant drawback of their paper. Raheel [6] proposed an 18-layer DCNN, and trained it on the pediatric CXR dataset following the dataset division of the original authors, i.e. 80% for training and 20% for test. The accuracy of their system (94.3%) was 1.6% higher than that of Kermany et al. [16], and the sensitivity was 99%, but the specificity was only 86%.

CNN is perceived as a black box that outputs a performance without relaying an understanding of that performance. Such lack of transparency can adversely affect the decision making. To improve this situation, Rajarman et al. [84] proposed a visual explanation of the prediction and activation of CNNs. They evaluated two models (VGG16 and CNN built from scratch), and visually presented their results. The lung boundaries were detected by an atlas-based detection algorithm [84], and the classification was performed by a DCNN. The VGG16 achieved a better learning and optimization outcome than customized CNN. The VGG16 improved the accuracy of distinguishing between pneumonia and normal images from 92.8% in Kermany et al. [16] to 96.2%. Meanwhile, the accuracy of distinguishing between viral and bacterial pneumonias was improved from 90.7% to 93.6%.

Abiyev et. al. [99] proposed customized CNN, competitive neural networks (CpNNs) with unsupervised learning, and backpropagation neural networks (BPNNs) with supervised learning for pathology detection from chest X-rays. Multiple experiments performed show that CpNN converges faster than CNN, however, the accuracy of the proposed CNN is higher than CpNN, BPNN as well as VGG16, VGG19 and CNN with GIST.

### 5.6. Segmentation by FCN and RCNN

Gu et al. [85] proposed a two-step method that classifies bacterial and viral pneumonias from chest radiographs. The first step extracts the lung regions from the x-ray images using an FCN. The second step classifies the lung regions into pediatric viral or bacterial pneumonia by a deep learning network with handcrafted features. The authors used the publicly available MC [40] and JSRT [42] datasets for segmentation, and data from the Guangzhou Women and Children’s Medical Center for binary classification. Feature extraction by transfer learning achieved higher accuracy (80.0% ± 2.02%) than other feature-extraction techniques. The performance was further improved by an ensemble of features (gray-level co-occurrence matrix, wavelets, histogram of oriented gradients, and DCNN features), although the improvement was slight (accuracy = 82.3% ± 0.14%).

Mask–RCNN [87] is a deep neural network for tasks such as segmentation. Jaiswal et al. [81] proposed Mask– RCNN with ResNet101 (and ResNet50) as a backbone detector for pneumonia detection, and trained it on the dataset of the Radiological Society of North America [88], which contains 30,000 annotated x-ray mages. When evaluated on the test set (the Society of Thoracic Radiology dataset [89]), Mask–RCNN outperformed several object detection techniques, such as YOLO3 and U-Net. The proposed model predicts the bounding box of each CXR, its label, and its masks with the respective class.

The method of Li et al. [90] identifies and localizes disease lesions in CXRs [1] under a limited amount of supervision. The proposed method accurately visualizes the location of the disease in the x-ray image, improving the disease interpretation. First, the image input is processed by Preact–Resnet (Resnet-v2) [91], which extracts the feature tensors of size *h’ = h/*32, *w’ = w/*32 *and c’ =* 2048. Here, *h, w*, and *c* are the height, width, and number of channels of the input image, respectively. The image is then divided into a *P* × *P* grid for predicting *K* possible disease types. As the recognition network, they applied FCN [92]. The image model was Resnet-v2-50, and the patch-slice size was selected from {12, 16, 20}. The CXR dataset contains 984 labeled bounding boxes for 880 images; the reaming 111,240 images are unannotated. The first evaluation was performed by five-fold cross-validation on 70% of the unannotated images and 70% of the annotated images. The second evaluation checked the effectiveness of the supervision provided by the bounding boxes. To this end, the proportion of unannotated bounding boxes was reduced from 80% to 0%. The authors showed that in some cases, the result of 40% (44496) unannotated images with 80% (704) annotated images outperformed the result of 80% (88; 892) unannotated images.

### 5.7. Ensemble methods

Chouhan et al. [93] proposed an ensemble of different state-of-the-art deep learning algorithms (Alexnet, Inception v3, Resnet, GoogleNet, and Densenet-121) for pneumonia detection in pediatric CXRs. Multiple experiments confirmed that the ensemble method outperformed other methods [16], achieving 96.4% accuracy and 99.0% sensitivity (versus an average of 92.8% accuracy and 93.2 sensitivity in previous methods). However, the precision (93.2%) was not appreciably changed from the previous methods.

Vijendran et al. [94] proposed a multi-layered method called Online Sequential Extreme Learning Machines (OSELM) for pneumonia detection in images of the MNIST dataset. The classification accuracy of multi-layered OSELM exceeded those of SVM and conventional extreme learning machines and achieved almost the same accuracy as traditional CNN within a shorter runtime. However, no additional performance metrics were provided in the paper.

Islam et al. [95] proposed ensemble DCCN models that accurately detect abnormalities (Cardiomegaly and Tuberculosis detection) in chest x-rays. They detected cardiomegaly events in the Indiana, JSRT, and Shenzhen datasets, detection, and achieved a 17% higher accuracy than single DCNN with 93.0% for Cardiomegaly detection and 90% for Tuberculosis detection.

Sirazitdinov et. al. [101] proposed ensemble of RatinaNet and Mask R-CNN for pneumonia detection and localization from dataset of 26,684 images from Kaggle Pneumonia [100] which achieved precision, recall and F-1 score of 0.75, 0.79 and 0.77, respectively.

### 5.8. Analyses of partial datasets with nonstandard divisions

Some researchers have performed non-standard divisions of partial datasets. Rahmat et al. [96] proposed Faster RCNN, which classifies pathological and normal CXRs. They selected a subset of 200 pathological and normal images from the Chest X-ray14 dataset. The accuracy of their Faster RCNN was 62.0%, higher than the judgement accuracies of a general practitioner and a medical student, but still very low because the data were highly imbalanced. The specificity (54.39%) was also low. Malygina et al. [62] proposed CycleGAN as a solution to the unbalanced data problem. They created samples using CycleGAN and classified them by DenseNet.

Varshni et al. [97] applied three deep learning algorithms (DenseNet, Resnet, Inception) as the feature extractor and SVM as the classifier of pneumonia signs in CXRs. As the Chest x-ray14 dataset [1] contains 1431 pneumonia images, they selected 1431 normal images to balance the dataset for binary classification. In a series of experiments with the appropriate parameters of the SVM classifier in DenseNet169, the AUC on the 573-image test dataset was 0.8002.

## 6. PERFORMANCE METRICS

The performance of a pneumonia detection system is evaluated by various performance metrics. The accuracy metric, which determines the correctness of the identified instances in both classes of a binary classification, must be supplemented by other metrics such as the precision, recall, *F*1 score, and *AUC*. The precision, recall, and *F*1 score are respectively given by Eqs. (1), (2) and (3) below. Table 1 shows the structure of a confusion matrix.

**TABLE 1:** PROVIDES A CLEAR DEPICTION OF CALCULATION THE PERFORMANCE METRICS.

| Actual Label | Prediction (Disease) | Prediction (Normal) |
| --- | --- | --- |
| Positive (Disease) | TP | FN |
| Negative (Normal) | FP | TN |

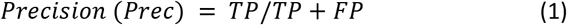

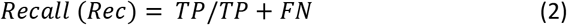

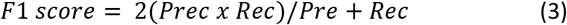

## 7. COMPARATIVE ANALYSIS AND DISCUSSION

In this section, we will briefly discuss and compare the relevant dataset and approaches for pneumonia detection. We will evaluate these approaches in terms of robustness and usability.

As can be seen in Table, there are multiple datasets available for the prescribed task. The most famous and most significant is to MIMIC CXR [37] and then the Chest X-rays-14 dataset [1]. However, these two datasets only contain fewer examples of pneumonia as compared to other classes, which makes them highly imbalanced. Although various techniques [43]-[71] have already been presented in the literature to tackle this problem, still only a few researchers have followed the techniques. In most cases, the authors have only utilized under-sampling and conclude the results with fewer images. The results re there for unreliable and cannot be exploited on an industrial scale. It can also be observed that most chest x-ray dataset did not contain any sample or pneumonia at all [24],[39],[41],[42],[98]. We suggest collecting massive scale data which should be balanced and should have around 1 million samples of pneumonia cases. As a temporary solution, the composite samples from the present dataset can also be exploited, but still, the combined image made only around five thousand images, which are far less for efficient and reliable detection.

As it can be seen from Table 3, that several author have presented significant amount of work in the pneumonia detection domain. Most of them have comparable accuracy, however the results are evaluated on small scale databases and cannot be utilized on a commercial scale. In the domain of traditional machine learning algorithms, Sousa et al. [98] have achieved 96% accuracy by exploiting NB with dimensionality reduction techniques. For traditional models, this much progress is worth noting and remarkable. By applying Transfer learning Kermany et al. [16] have achieved 90.7% accuracy which is lower as compared to Sousa et al. [98]’s performance. Chouhan et al. [93] have achieved 96.2% accuracy by employing assemble method which contains a major part of DCNN, and the computational resources required for such type of models are huge. Therefore, the current best performance with less computational requirement is provided by Sousa et al. [98]. However, if the computational power is not constrained and performance is the only requirement, than the work of Chouhan et al. [93] can also be considered as remarkable. Besides, there are several other works which have provided <90% accuracy for pneumonia detection such as Rajarman et al. [96],, Stephan et al. [5], and Raheel [6].

**Table 2:**
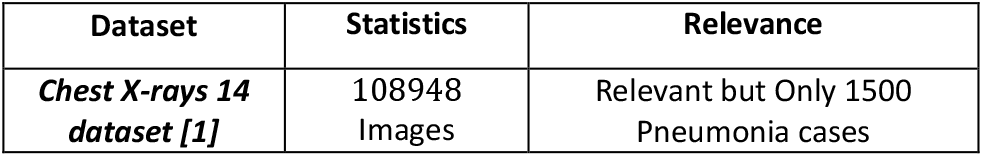
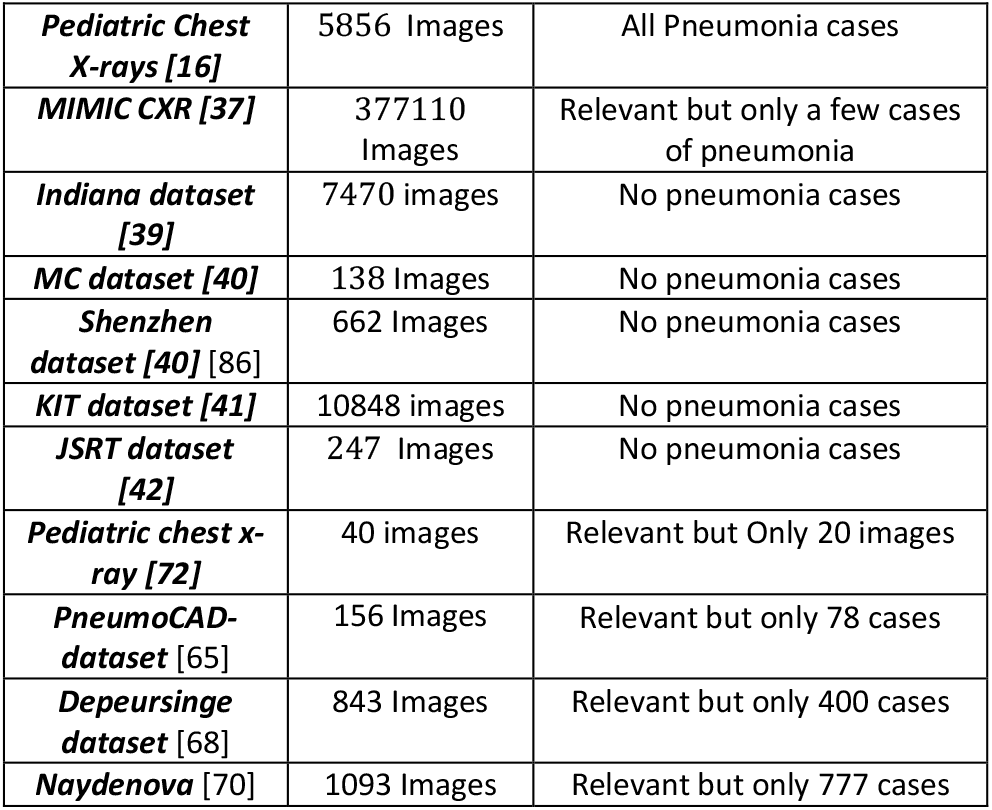
Comparison of datasets comprised of chest x-ray w.r.t detection of pneumonia

**Table 3:**
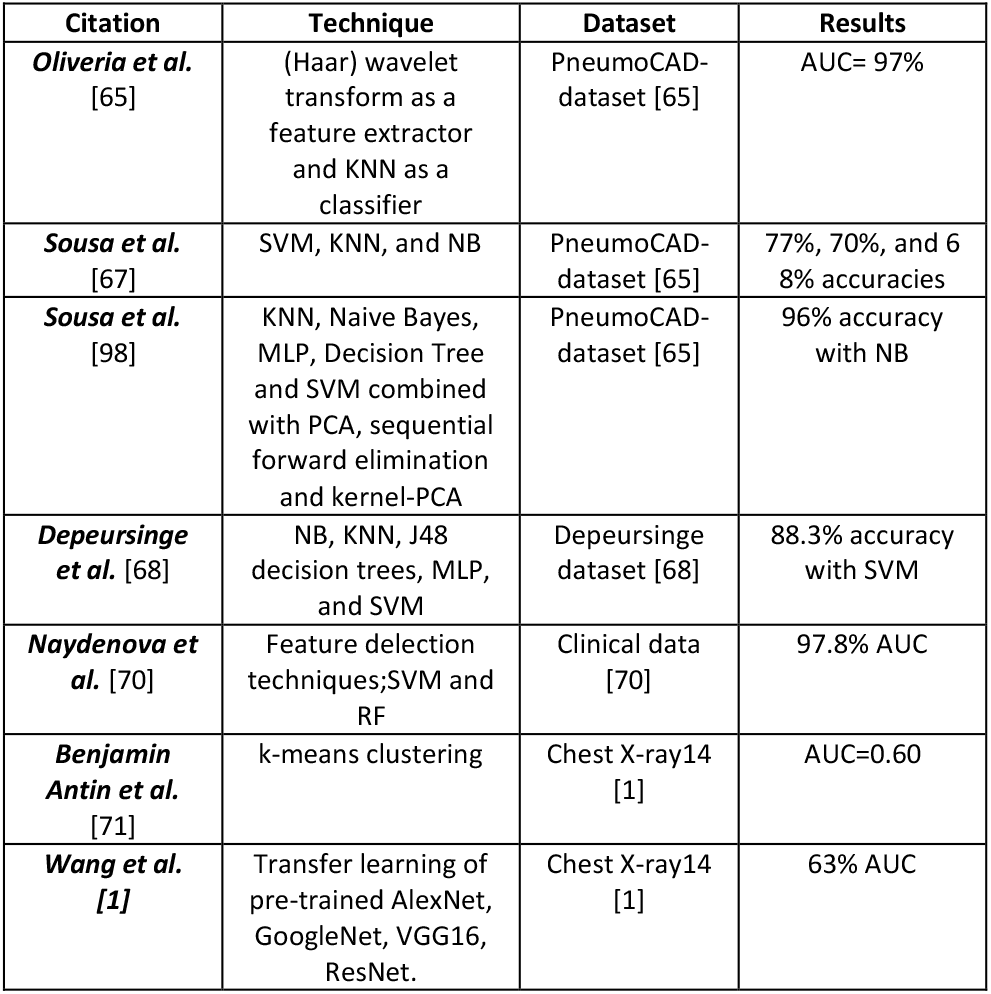
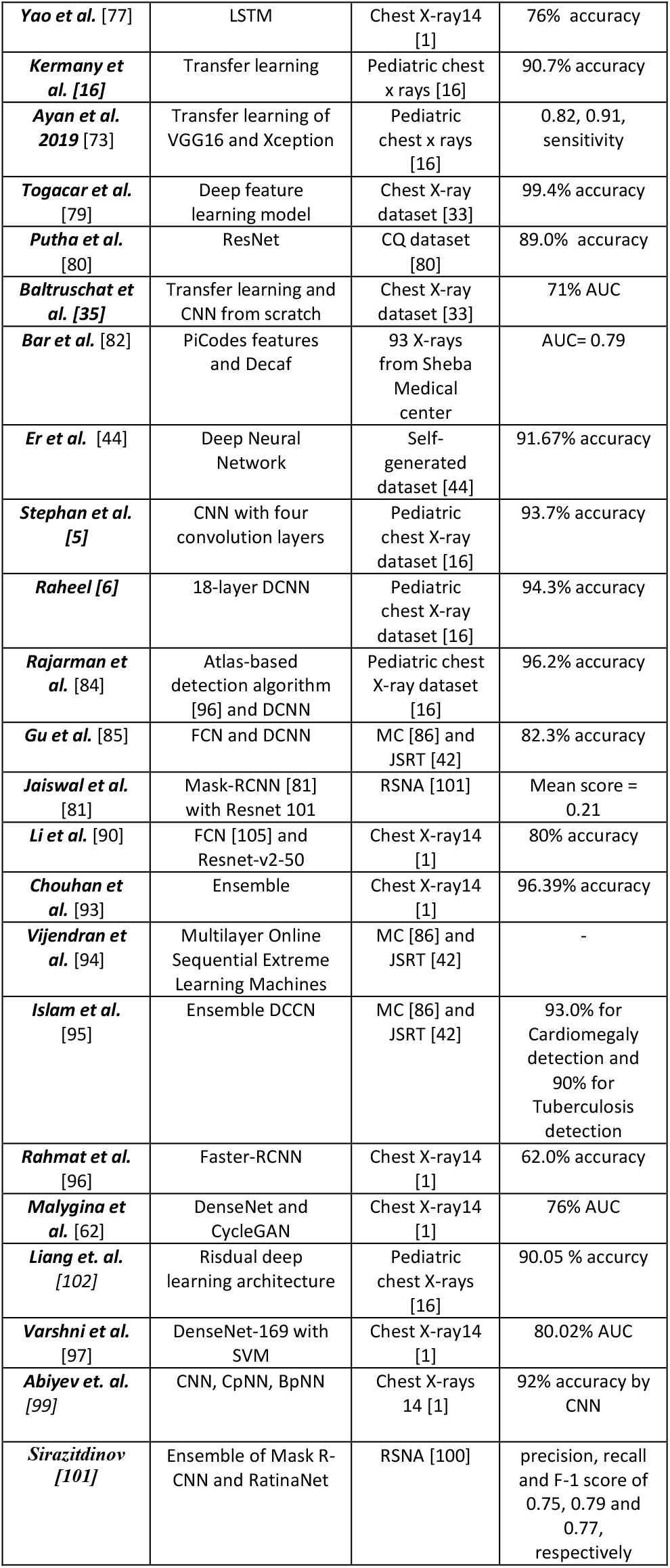
Comparison of pneumonia detection techniques

## 8. CONCLUSION

The use of chest radiography has a vital role in the examination and diagnosis of chest related diseases, which made the automatic detection one of the hot topics in computer vision as medical imaging research. Thus, many algorithms using various techniques are available from the research community. However, there is a lack of available literature that summarizes all the current available practices, so one can visualize what methods to choose as a real-time perspective, which are the available datasets, and what are the currently achieved results in this domain.

This paper presented the overview of current literature on the topic of pneumonia identification by using chest x-ray, it summarizes the topic and provides the analysis of present algorithms in terms of usability, goodness factor, and computational complexity associated with these techniques. We have observed that there are multiple datasets available for the prescribed task.

However, most of them are highly imbalanced and only a few researchers have followed the balancing techniques. In most cases, the authors have only utilized under-sampling and conclude the results with fewer images. The results are there for unreliable and cannot be exploited on an industrial scale. We suggested collecting a massive scale data which should be balanced and should have around 1 million samples of pneumonia cases.

Similarly, several authors have presented significant amount of work in the pneumonia detection domain. The current best performance with less computational requirement is provided by Sousa et. al. [98]. However, if the computational power is not constrained and performance is the only requirement, than the work of Chouhan et al. [93] can also be considered as remarkable.

## Data Availability

The data that support the findings of this study are available on request from the corresponding authors.

